# Molecular profiling of primary renal diffuse large B-cell lymphoma unravels a proclivity for immune-privileged organ-tropism

**DOI:** 10.1101/2024.07.16.24309727

**Authors:** Axel Künstner, Vera von Kopylow, Philipp Lohneis, Matthias Kümmel, Hanno M. Witte, Lorenz Bastian, Veronica Bernard, Stephanie Stölting, Kathrin Kusch, Manuela Krokowski, Nikolas von Bubnoff, Konrad Steinestel, Annette Arndt, Hartmut Merz, Hauke Busch, Alfred C. Feller, Niklas Gebauer

## Abstract

Primary renal manifestations of diffuse large B-cell lymphoma (PR-DLBCL) represent an exceptionally rare variant of the most common type of non-Hodgkin lymphoma (NHL). Insights into PR-DLBCL pathogenesis have been limited to small case series and methodologically limited approaches. The mechanisms driving lymphomagenesis within an organ lacking an intrinsic lymphatic niche and its proclivity for dissemination to immune-privileged sites, including testes and central nervous system, remain poorly understood. To decode the genetic and transcriptional framework of PR-DLBCL, we utilized whole exome sequencing, array-based somatic copy number alterations analysis, and RNA sequencing. Hereby we characterize the most extensive cohort of PR-DLBCL published, comprising 34 samples from 30 patients. Despite significant mutational heterogeneity with a broad distribution among molecular clusters, we observed a strong unifying enrichment in deleterious MHC class I and II aberrations and loss of *CDKN2A* at a frequency similar to primary large B-cell lymphoma of immune privileged sites (IP-LBCL) alongside significant transcriptional deregulation of interferon signaling and MYC targets in MHC class I-deficient cases.

Our integrative assessment of PR-DLBCL biology expands the molecular understanding of this rare variant including similarities with IP-LBCL as an intriguing explanation for its clinical behavior and tropism. Our observations may inform future risk-adapted therapeutic approaches.

## Introduction

Diffuse large B-cell lymphoma (DLBCL) is the most common type of non-Hodgkin lymphoma (NHL) and accounts for 30 – 40% of newly diagnosed lymphomas ^1^. Extranodal manifestations in both the adrenal glands and the kidneys constitute long-standing risk factors for developing simultaneous or secondary CNS manifestations ^2^. Secondary renal involvement is recurrently observed in advanced-stage non-Hodgkin lymphoma (NHL), whereas primary renal (PR-) NHL manifestations are notably less frequent, leading to skepticism regarding the existence of primary renal lymphoma for an extended period ^2–6^. Population-based datasets suggest an age-adjusted incidence of 0.035/100,000 with an increasing tendency, a median age of approximately 70 years at diagnosis, a male predominance, and mostly unilateral tumors. Among PR-NHL, DLBCL appears to be the most common histology followed by marginal zone (MZL) and follicular lymphoma (FL) ^7, 8^. Clinical differentiation of PR-NHL from other renal malignancies including renal cell carcinoma poses a long-standing challenge, which led to a substantial number of PR-NHL patients undergoing curatively intended resection ^9^. In recent years, the introduction of FDG-PET-CTs into routine clinical practice has enabled a more reliable characterization, particularly in patients with more aggressive PR-NHL^10^.

Insufficient characterization of pathogenetic mechanisms in indolent PR-DLBCL derives from limited data, primarily comprised of small case series and a lack of in-depth molecular characterization of patients and samples ^11^. Molecular drivers of lymphomas originating from organs lacking an intrinsic lymphatic niche, such as the kidney, remain widely elusive apart from individual case studies ^12^. To elucidate the genetic and transcriptional landscape and drivers of PR-DLBCL, we conducted a comprehensive investigation employing whole exome sequencing (WES), array-based analysis of somatic copy number alterations (SCNA), and RNA sequencing (RNA-seq) to unravel both molecular properties of this rare entity as well as the cellular composition of its tumor microenvironment (TME).

## Materials and methods

### Case selection and clinicopathological assessment

For this retrospective analysis, we reviewed our institutional archive for cases of histologically confirmed primary renal DLBCL between January 2001 and December 2021. Only such cases without concurrent non-renal lymphoma manifestations at diagnosis were included in the study, which led to a large subset of samples obtained by complete renal resection under the suspicion of non-metastatic renal cell carcinoma (22/34 samples; 65%). The study was approved by the ethics committee of the University of Lübeck (reference-no 18-356) and conducted per the declaration of Helsinki. Patients provided written informed consent regarding routine diagnostic and academic assessment, including genomic studies. Histopathological work-up was performed as described and diagnosis was confirmed following the 5^th^ edition WHO classification of hematopoietic and lymphoid tumors and the ICC criteria and yielded 34 cases of PR-DLBCL for which sufficient formalin-fixed paraffin-embedded (FFPE) tissue samples were available for subsequent molecular analysis ^1, 13, 14^.

### Delineation of the molecular composition of the study cohort

Extraction of nucleic acids, WES and RNA-seq alongside detection of Somatic Copy Number Alterations employing the OncoScan CNV array (ThermoFisher was performed as described, and Raw fastq files have been added to the European Genome-phenome archive (EGA) under the previous accession number EGA50000000386 ^13, 15^. OncoScan Array data has been deposited in Gene Expression Omnibus (GEO) under accession number GSE270422.

### Whole exome data processing and variant calling

The raw sequencing data in FASTQ format was processed using the NFORE workflow (NEXTFLOW v23.10.1) SAREK (v3.3.2) for variant calling against the GRCh38 reference genome ^16, 17^. Initially, reads were trimmed for adapter sequences and quality using FASTP (v0.23.4), followed by alignment with BWA MEM2 (v2.2.1) ^18, 19^. Subsequent steps included mate-pair information correction, removal of PCR duplicates, and base quality recalibration utilizing PICARD TOOLS, GATK (v4.4.0.0), and dbSNP v146 ^20, 21^. Finally, variant calling was executed in tumor-only mode using MUTECT2 with GNOMAD (r2.1.1) as the reference for known germline variants ^22, 23^. Variants were left aligned (GATK LEFTALIGNANDTRIMVARIANTS) and only variants with a PASS filter flag were kept for variant annotation using VARIANT EFFECT PREDICTOR (VEP v111, GRCh38; adding CADD v1.6, dbNSFP v4.1a, and GNOMAD r3.0 as additional annotations) ^24–26^. Next, annotations were converted into *MAF* format using VCF2MAF (V1.6.21) (DOI:10.5281/ZENODO.593251); coverage was extracted directly from the vcf INFO field. Potential FFPE artifacts in the variant data were identified using two approaches. First, strand orientation biases (mutations just found on one strand, F1R2 or F2R1) were detected by SOBDETECTOR (v1.0.4) and potential artifacts were removed ^27^. Next, a classifier detecting the origin of mutation in formalin-fixation paraffin-embedding (FFPE) samples was applied (R-package EXCERNO v0.1.0) ^28^. Variants annotated with an FFPE-like signature were removed. Potential germline variants in the data were removed using GATKs 1000g Panel of Normals. The top 20 frequently mutated genes (FLAGS)^29^ were removed from further analysis and the remaining somatic variants were filtered as follows: minimum coverage of 50, minimum alternative allele coverage of 5, minimum variant allele frequency of 10%, and only variants with a frequency < 0.1% in 1000 genomes, GNOMAD, or ExAC were considered for subsequent downstream analysis; variants were required to have a COSMIC or dbSNP ID. High-impact variants (CADD score > 20) in candidate genes (see below) were filtered as such that minimum coverage of 10, minimum alternative coverage of 4, and minimum variant allele frequency of 10% was required. Candidate genes were defined as follows: (a) listed as tumor suppressor or oncogene according to Vogelstein et al. ^30^, (b) genes from the Lymphgen algorithm, (c) DLBCL genes described by Chapuy *et al.*, (d) MHC genes according to Harmonizome (v3.0) ^31–33^. A brief graphical representation of the bioinformatics workflow for WES data processing is provided in **Supplementary Figure S1**.

### Detection of Somatic Copy Number Alterations

Somatic copy number alterations were detected using OncoScan CNV assays (ThermoFisher). Raw data (CEL files) were processed using the EACON package (v0.3.6-2) with SEQUENZA as segmentation algorithm ^34^. L2R files in CBS format were used as input for GISTIC (v2.0.22) to identify regions significantly amplified or deleted across all samples (confidence level 0.90, focal length cutoff 0.5, q-value threshold 0.1) ^34, 35^.

For details on transcriptome data quantification, fusion detection, statistical analysis, and pseudonymization, please see **Supplementary Materials and Methods**.

## Results

### Clinicopathological Characteristics and Cell-of-Origin in PR-DLBCL

We collected 34 PR-DLBCL samples from 30 patients with sufficient FFPE tissue samples for in-depth molecular characterization (**Figure 1A**). The median age of the study group was 66.5 years, 13 patients were female and 17 were male. No iatrogenic immunosuppression, immunodeficiency, and history or current presence of other lymphomas were reported in any of the patients. In particular, only patients without suspicion of concurrent lymphoma manifestations were eligible for the study. This is mirrored in the exceptionally high frequency of cases that underwent surgical resection of the kidney (22/34 cases; 65%). Baseline clinicopathological data are summarized in **Supplementary Table 1**.

**Figure 1.**
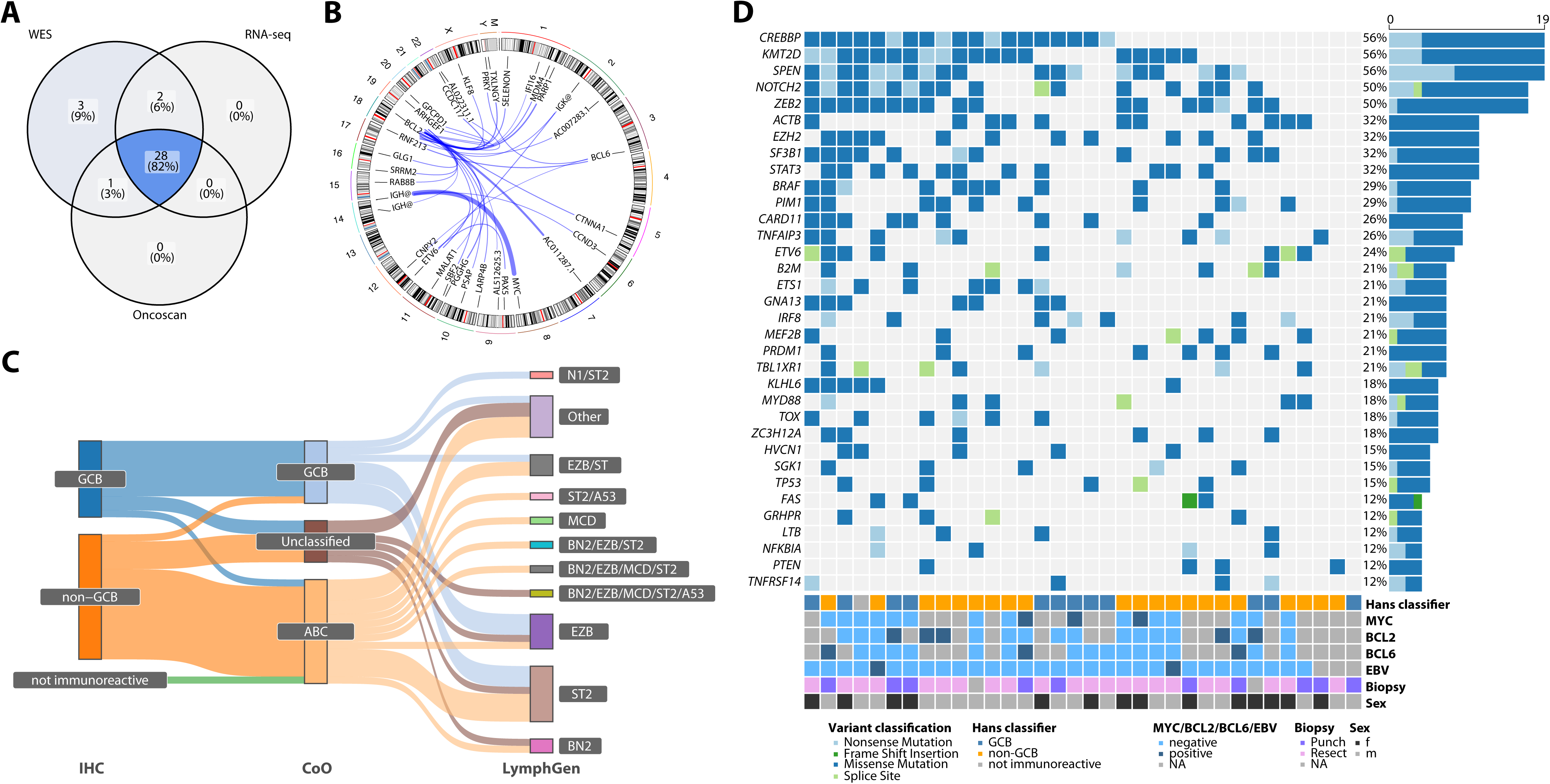
Molecular landscape of PR-DLBCL. **A** Venn diagram depicting the number of samples for which WES (n = 34), RNA-seq (n = 30), and OncoScan arrays (n = 29) were successfully conducted. **B** Fusion identified by RNA-sequencing in PR-DLBCL samples (beyond *MYC*/*BCL2*/*BCL6* rearrangements identified in cases studied by FISH) displayed by their genomic location. **C** Sankey plot illustrating the distribution of cases of PR-DLBCL into the categories GCB/non-GCB by immunohistochemistry (IHC) according to the algorithm proposed by Hans *et al.*^38^ as well as by Cell-of-origin (CoO) classified according to gene-expression profiling ^39^ derived from RNA-seq and lastly according to molecular clusters drawn from the LymphGen algorithm, integrating data from WES, RNA-seq, Oncoscans, and FISH (*BCL2*/*BCL6*/*MYC*) ^31^, depicting the significant degree of molecular heterogeneity. **D** Oncoplot displaying putative driver genes and the number of samples harboring mutations in a given gene (right bar). Mutation types are color-coded, and covariates, including sex, CoO, FISH results for BCL2/BCL6/MYC, and type of biopsy (punch/needle core vs. open resection/nephrectomy) are shown below for each sample.

Structural variants in *BCL2*, *BCL6*, and *MYC* were assessed successfully in 19, 18, and 17 patients by FISH break-apart probes. In cases without sufficient tissue for FISH but with RNA-seq data, we screened for high-confidence *BCL2/BCL6/MYC* fusions (**Figure 1 B**). Here, we observed aberrations in five (26 %), two (11 %), and three (18%) cases respectively, indicative of a slightly elevated frequency of these aberrations in comparison to DLBCL (NOS) of other primary manifestations. In addition, we observed two cases with ETV6-Ig fusions, previously described in PCNSL another case of an ETV6:PAX5 fusion respectively, previously described to lead to similar ETV6 activation in B-cell precursor acute lymphoblastic leukemia ^36, 37^. Manually curated oncogenic fusions derived from RNA-seq data are summarized in **Supplementary Table 2**. By immunohistochemistry according to the Hans *et al.* algorithm in all samples with sufficient tissue available after DNA/RNA extraction, we observed the majority of cases to be of non-GCB type (18/30; 60%), whereas only a minor subset was of GCB type (11/30; 37%), and one case could not be classified due to insufficient immunoreactivity ^38^. Only one case was EBV-associated (>50% positive tumor cells by EBER-ISH). To further unravel the cell of origin, we performed RNA-seq on samples from 30 patients and applied the approach proposed by Wright *et al.* and found 15/30 cases (50%) to be of ABC-subtype. In comparison, 9/30 (30%) were GCB-DLBCL and six remaining cases (20%) remained unclassifiable ^31, 39^ (**Figure 1 C**).

### The mutational landscape of PR-DLBCL

To elucidate the mutational landscape of PR-DLBCL and to screen for mutational drivers of the subgroup’s peculiar tropism, we performed WES on FFPE tissue samples from 34 patients (**Figure 1 D**).

Adhering to stringent variant filtering, outlined above, oncogenic mutations were identified in all PR-DLBCL cases. In total 3,178 presumably deleterious mutations, affecting 1,388 genes were observed. Additionally, 25,457 somatic copy number aberrations (SCNAs) were detected. SNVs and indels comprised 11% of these mutations, of which 2,673 were missense (84%), 333 nonsense (10%) and 70 indel mutations (2%). Mutations affecting splice sites represented 3% of estimated somatic mutations and one of the observed mutations was a non-stop mutation (**Supplementary Table 3**). At an intermediate-low median tumor mutational burden (TMB) of 2.45 mutations/Mb (range 0.18 – 5.76), we observed no cases of microsatellite instability as could be expected in aggressive B-cell lymphomas. Variants per variant class as well as variants per sample including information on variant subtypes are provided in **Supplementary Figure S2**.

The most commonly mutated genes included chromatin-remodeling genes *CREBBP* and *KMT2D* both in 56% of cases with > 20% nonsense mutations, followed by NOTCH signaling-associated genes *SPEN* and *NOTCH2* (both 50%) with nonsense mutations in 47% and 18% respectively. Overall, we observed an enrichment in IL6/JAK/STAT3 signaling mutations including *STAT3*, *PIM1*, and *MYD88*. While *STAT3* mutations were distributed throughout the entire gene, we observed mutational impairment of pre-described hotspots in *MYD88* and *PIM1* (all restricted to the kinase domain) (**Figure 2**). Other biological processes targeted by deleterious mutations in PR-DLBCL included the epigenetic regulation of gene expression (*SF3B1*, *KMT2D*, and *EZH2*), as well as B-cell receptor and RTK-RAS signaling (*CARD11*, *PRDM1*, and *BRAF*). The distribution of mutation onto the candidate driver genes referenced above is displayed in **Figure 2**. In a more global approach, we assessed the impairment of key biological processes through an enrichment analysis of mutations against previously described oncogenetic gene sets and against the IL6/JAK/STAT3 signaling pathway (**Figure 3**) ^40^. Hereby we uncovered a relatively wide distribution within a given set (**Figure 3A**). Intriguingly, the most commonly affected gene sets included NOTCH, RTK-RAS, and IL6/JAK/STAT signaling at 97%, 94%, and 91%, respectively (**Figure 3 B, C**). Beyond this, we observed mutational impairment of PI3K signaling in 79% of cases, suggesting a significant role in PR-DLBCL pathogenesis. Correlating information on cell of origin and mutational patterns, we observed a statistically significant enrichment in mutations affecting *TNFAIP3* and *PIM1* among non-GCB cases. *BCL2* rearranged cases showed significantly more *DNMT1* and *SMARCA4* mutations (**Supplementary Figure S3**). We additionally screened for recurrent mutations in canonical tumor suppressor genes (TSGs) and found deleterious mutations in 33/34 (97%) cases. Among the most common targets, in addition to *CREBBP* and *KMT2D,* we observed mutations in *TP53* in 15% of cases.

**Figure 2.**
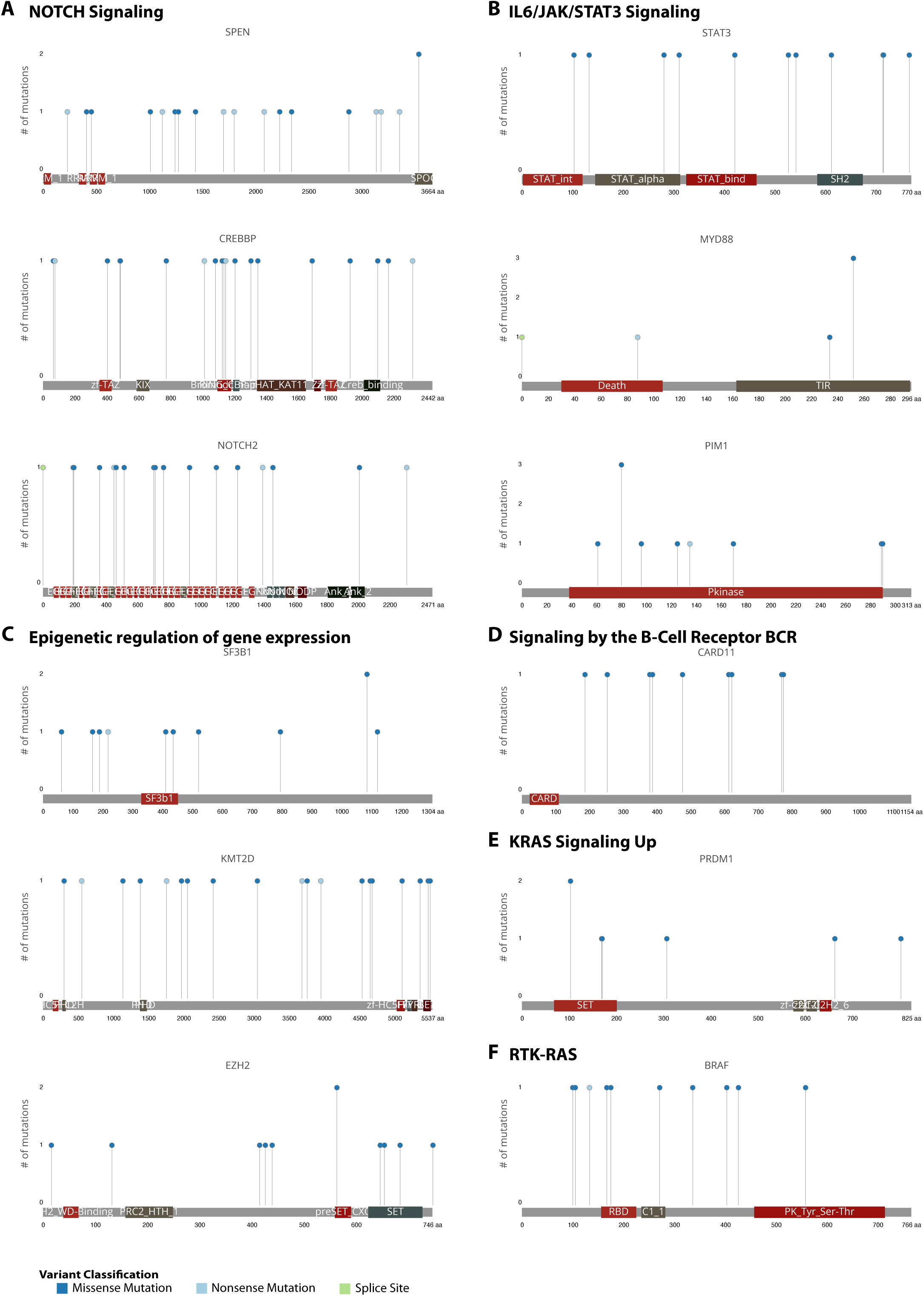
Recurrently mutated putative drivers of PR-DLBCL. Lollipop plots illustrating types and location of mutations, grouped by affected signaling pathway and/or biological function. Functional domains are depicted as a frame of reference and the lollipops’ height indicates the frequency with which a given mutation occurred in our cohort.

**Figure 3.**
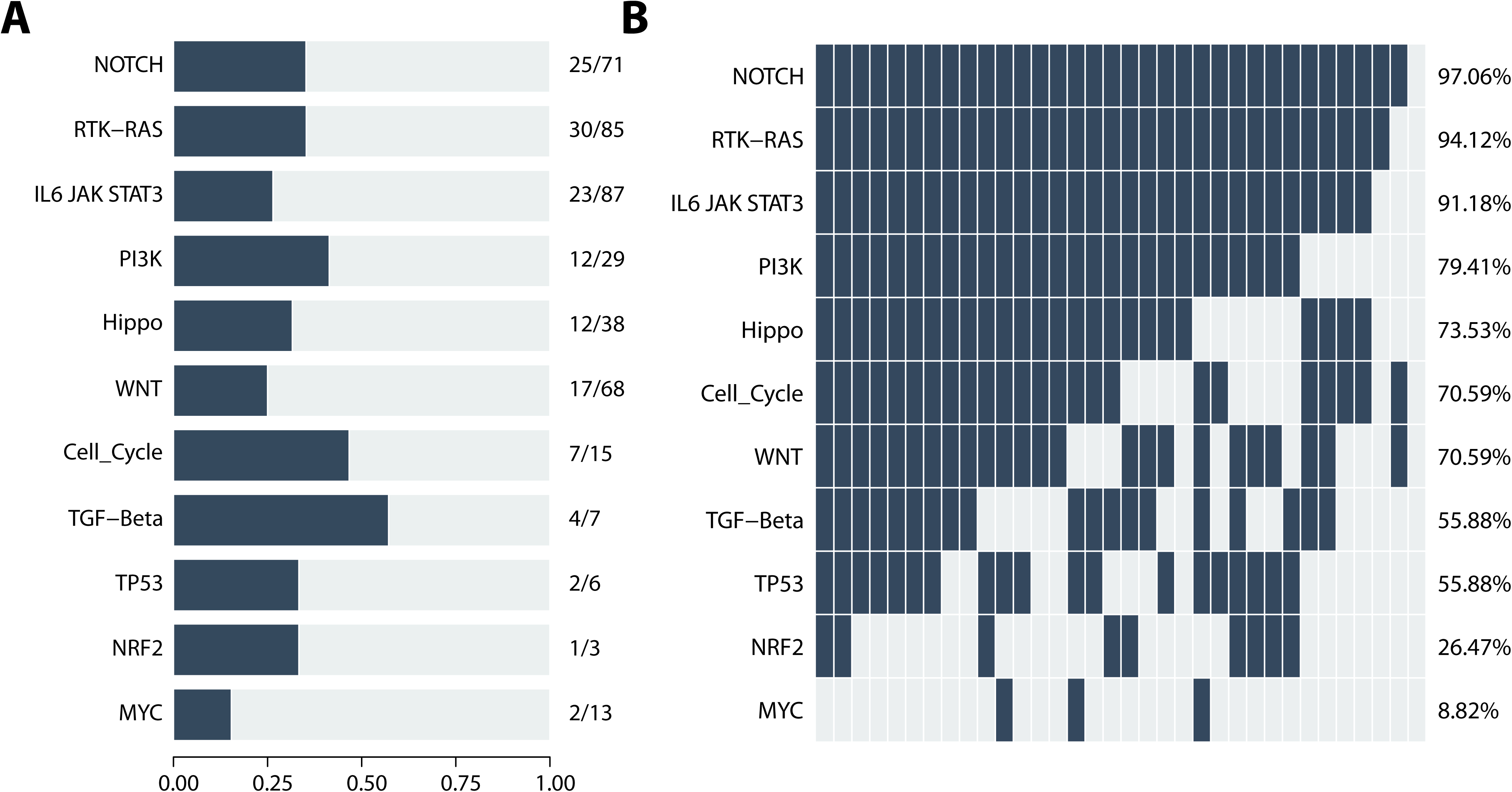
Functional implication of mutations in PR-DLBCL. **A** Fraction of a given pathway, biological function, or gene set impaired by mutations is depicted as the number of affected genes vs. the number of genes in a given set. **B** Distribution of mutational impairment of gene sets per sample, including fraction of samples harboring one or more mutations in a given gene set.

### Genome-wide analysis of somatic copy-number variations in primary renal DLBCL

To unravel the genome-wide landscape of SCNA in PR-DLBCL, we analyzed 29 samples successfully on the Illumina OncoScan array and subsequently employed GISTIC (v2.0.22) to identify regions significantly amplified or deleted across all samples. Arguably the most striking observation was a broad arm-level deletion spanning a large region on chromosome 6 in 48% of evaluable cases (**Figure 4A**). Candidate genes, previously implicated in lymphoma pathogenesis located in this vast region included *PRDM1*, *NFKBIE*, *HLA-A*, *HLA-B*, and *ARID1B*. Beyond this aberration, the genome-wide CNV landscape of PR-DLBCL is dominated by CNVs affecting 3p12.1 and leading to copy number losses of VHL, SETD2, and others (17%), 9p21.3 encompassing the region encoding for TSG *CDKN2A* (38%), as well as the *NOTCH1* region at 9q34.3 (28%). Focal amplifications were observed at 1q23.1, 3p12.3, 3q29, and 22q11.23. Candidate genes among significant deletions and amplifications are provided in **Supplementary Table 4** and **Figure 4B**. To assess the characteristics of PR-DLBCL in contrast to transcriptionally defined subtypes of DLBCL (ABC/GCB) and primary CNS lymphoma (PCNSL) as well as primary testicular large B-cell lymphoma (PTLBL) together representing the subgroup of primary large B-cell lymphomas of immune-privileged sites. We conducted a comparative analysis drawing CNV data from two previously published large-scale genome-wide studies (**Figure 4 C**) ^41, 42^. Apart from a focal 3q29 amplification, none of the previously reported copy number gains were observed at a significant level in our cohort. At the same time and of particular interest, we observed strong similarities between PR-DLBCL, PCNSL, and PTLBL regarding deletions at 6p21.33, 6q21, and 6q23.3. Candidate gene deletions previously implicated in post-germinal differentiation and strong CNS tropism located at these loci include *HLA-B* and *PRDM1*. Moreover, we identified the above-mentioned *CDKN2A* deletions in PR-DLBCL at a frequency more similar to PCNSL/PTLBL than an all-comer DLBCL cohort. Additionally, we observed a significant impact of SCNAs on gene-expression profiles which is shown exemplarily for TSGs (**Supplementary Figure S4**).

**Figure 4.**
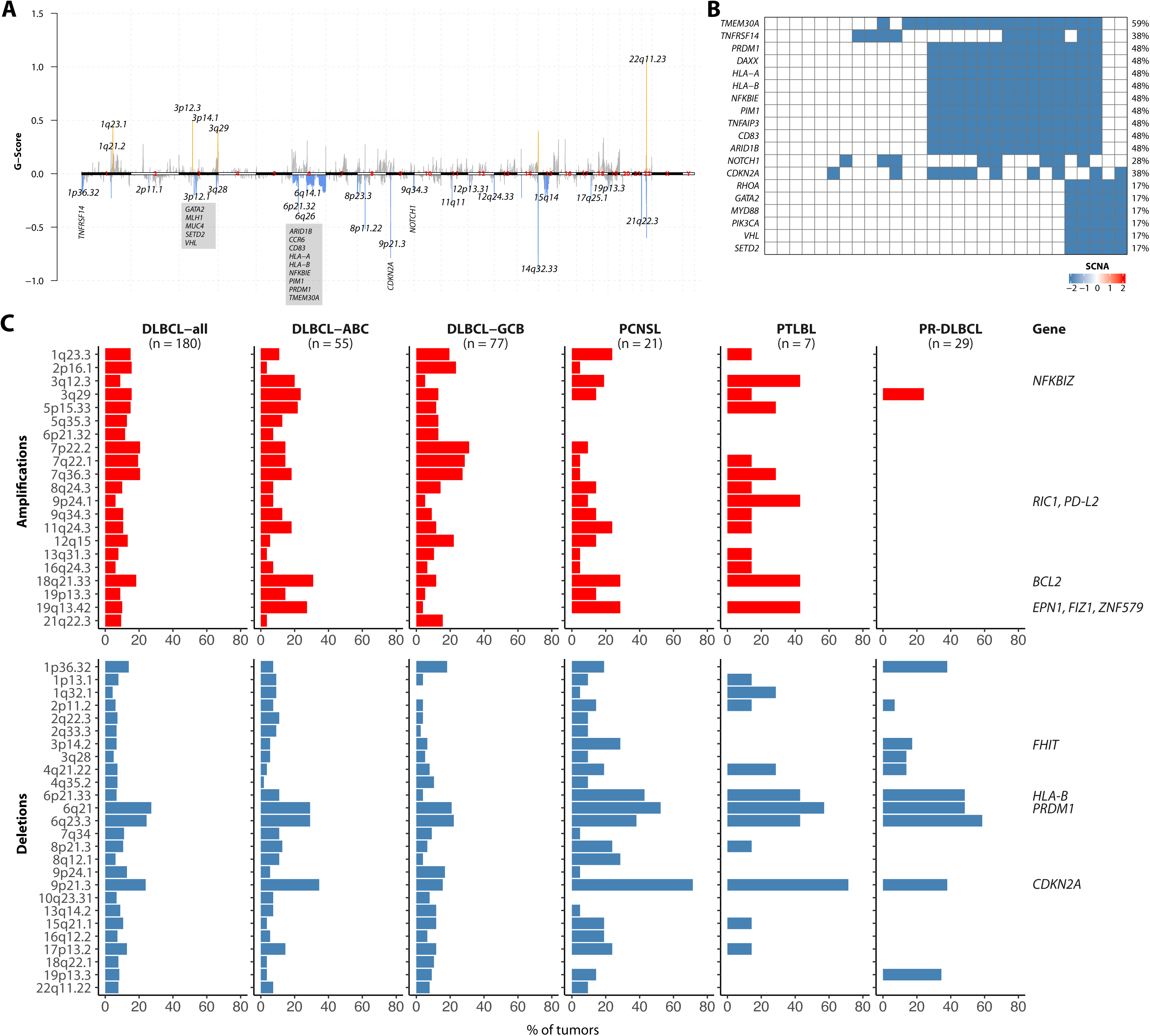
Somatic copy-number aberrations in PR-DLBCL compared with other subtypes of DLBCL including IP-LBCL. **A** Location of SCNAs along the genome and GISTIC G-scores (G = Frequency × Amplitude; red bars denote gains and blue bars losses; gene names refer to affected oncogenes and tumor suppressor genes within identified regions). **B** Tumor suppressor genes affected by copy number losses. **C** Amplifications and deletions at recurrently affected loci in an all-comer cohort of DLBCL (first column), DLBCL of ABC-as well as GCB-subtype (second and third column) and IP-LBCL with primary CNS or primary testicular manifestation (column four and five) compared to primary renal DLBCL (column six).

### Molecular clusters in primary renal DLBCL

Building on our observations from WES, CNV analysis, RNA-seq, alongside structural variants identified by FISH for *BCL2*, *BCL6*, and *MYC*, we performed an integrative analysis on our entire cohort of PR-DLBCL samples to achieve an allocation to the previously defined molecular clusters (A53, N1, BN2, EZB, and MCD) based on the probabilistic LymphGen 2.0 classifier ^31^. Molecular data for an integrative analysis of sufficient sensitivity and specificity was available in 30 cases. Unlike previous observations in PCNSL/PTLBL, only one case resembled MCD-DLBCL. The most common molecular subtypes were ST2 (eight cases, 27%) and EZB (five cases, 17%). Of note, no double- or triple-hit lymphomas including MYC+ EZB cases were observed. Only two cases were BN2-DLBCL (7%). While a significant overlap between signatures with compound-cluster allocation was observed in eight cases, six cases were found to exhibit a constellation of findings leading to a classification as “other” (**Figure 1 D** and **Supplementary Table 5**).

### Immune-escape mechanisms in primary renal DLBCL as a driver of tropism

Following the characterization of the mutational profiles and molecular clusters that displayed the significant heterogeneity outlined above, we sought to understand the underlying mechanism behind the exceptional renal tropism in our present cohort. Building on our observation of a significant overlap between PR-DLBCL, PCNSL, and PTLBL, in particular regarding copy number losses affecting HLA-B as a hallmark gene of MHC Class I, we sought to assess the extent to which the clinical behavior of PR-DLBCL might be explained by immune escape strategies. Integrating all available datasets, we identified deleterious MHC Class I lesions in 72% of cases, including HLA-A and HLA-B deletions in 48% and deleterious B2M mutations in 21% of evaluable patients (**Figure 5A, B**). An additional fraction of 33% of patients exhibited deleterious aberrations within the MHC class II machinery and 60% of patients additionally exhibited molecular hits in immediate interaction partners of MHC class I or II required for adequate immune response including del 19p13.3 and mutations affecting CD70, CIITA, and CD58 (**Figure 5A, C**).

**Figure 5.**
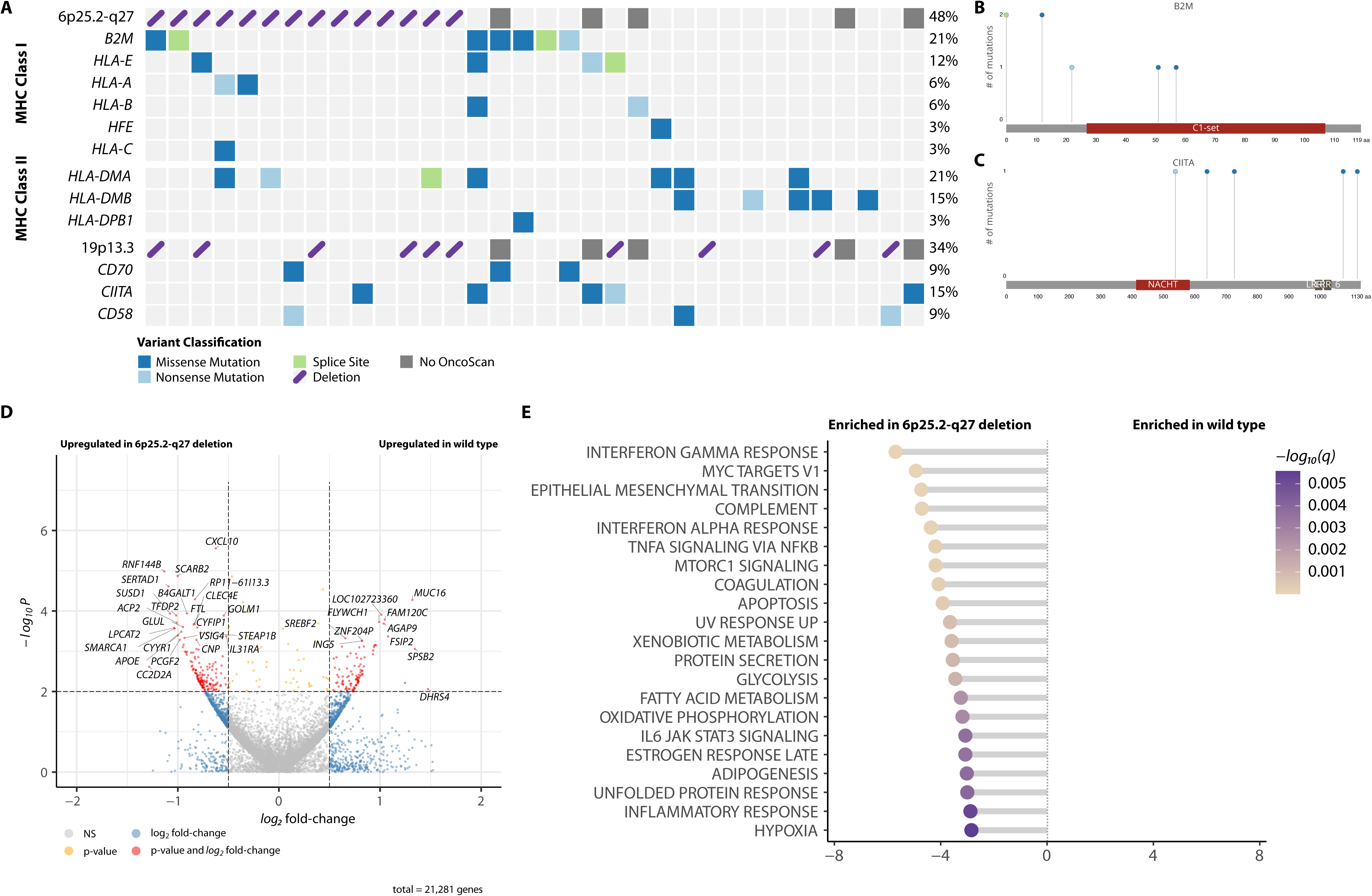
Immune escape is the predominant molecular feature of PR-DLBCL. **A** Impairment of the MHC class I and II apparatus as well as its immediate interaction partners by mutations and/or SCNAs. **B, C** Lollipop plots illustrating types and location of mutations in *B2M* and *CIITA* as recurrently mutated genes associated with immune escape in PR-DLBCL. Functional domains are depicted as a frame of reference and the lollipops’ height indicates the frequency with which a given mutation occurred in our cohort. **D** Volcano plot illustrating results of a genome-wide screen for significantly deregulated genes by the presence or absence of a large-scale deletion on chromosome 6 leading to strong MHC class I impairment as the most recurrent and striking feature in PR-DLBCL. **E** Gene-set enrichment analysis depicting functional implications of deregulated gene expression between cases with and without large-scale MHC class I deletions.

Having identified the large-scale 6p25.2-q27 deletion outlined above as a potential hallmark of aggressive B-cell lymphomas with renal tropism, we went on to assess the transcriptional profile of cases carrying said aberration and observed a significantly elevated expression of several candidate genes (**Figure 5D** and **Supplementary Table 6**) including *CXCL10* as well as strong enrichment for interferon alpha and gamma response, MYC targets, JAK/STAT signaling, and others (**Figure 5E** and **Supplementary Table 7**). In particular, we found downstream NFkB activation via TNFalpha signaling. In addition, several metabolic processes including glycolysis, fatty acid metabolism, and oxidative phosphorylation appear to be particularly engaged compared to non-MHC-class I-deleted cases.

Subsequent analysis of our cohort deconvoluting cell states and their assembly into communities of co-association patterns in the form of lymphoma ecotypes (LE) employing the lymphoma EcoTyper revealed a relatively diverse composition of both the tumor as well as its microenvironment in PR-DLBCL ^43^. Despite the limited number of cases, we observed a trend towards enrichment in prognostically adverse ABC-DLBCL-associated lymphoma ecotypes (LE) 2 and 4 dominated by B-cells, plasma cells, and dendritic cells as well as NK cells, CD4 and CD8 positive T-cells alongside regulatory T-cells, respectively in del 6p25.2-q27 cases. Moreover, a trend towards enrichment of the T follicular helper cells dominated LE5 was observed in non-del 6p25.2-q27 cases ^43^. The distribution of ecotypes on cases is depicted in **Figure 6** (**Supplementary Table 8**).

**Figure 6.**
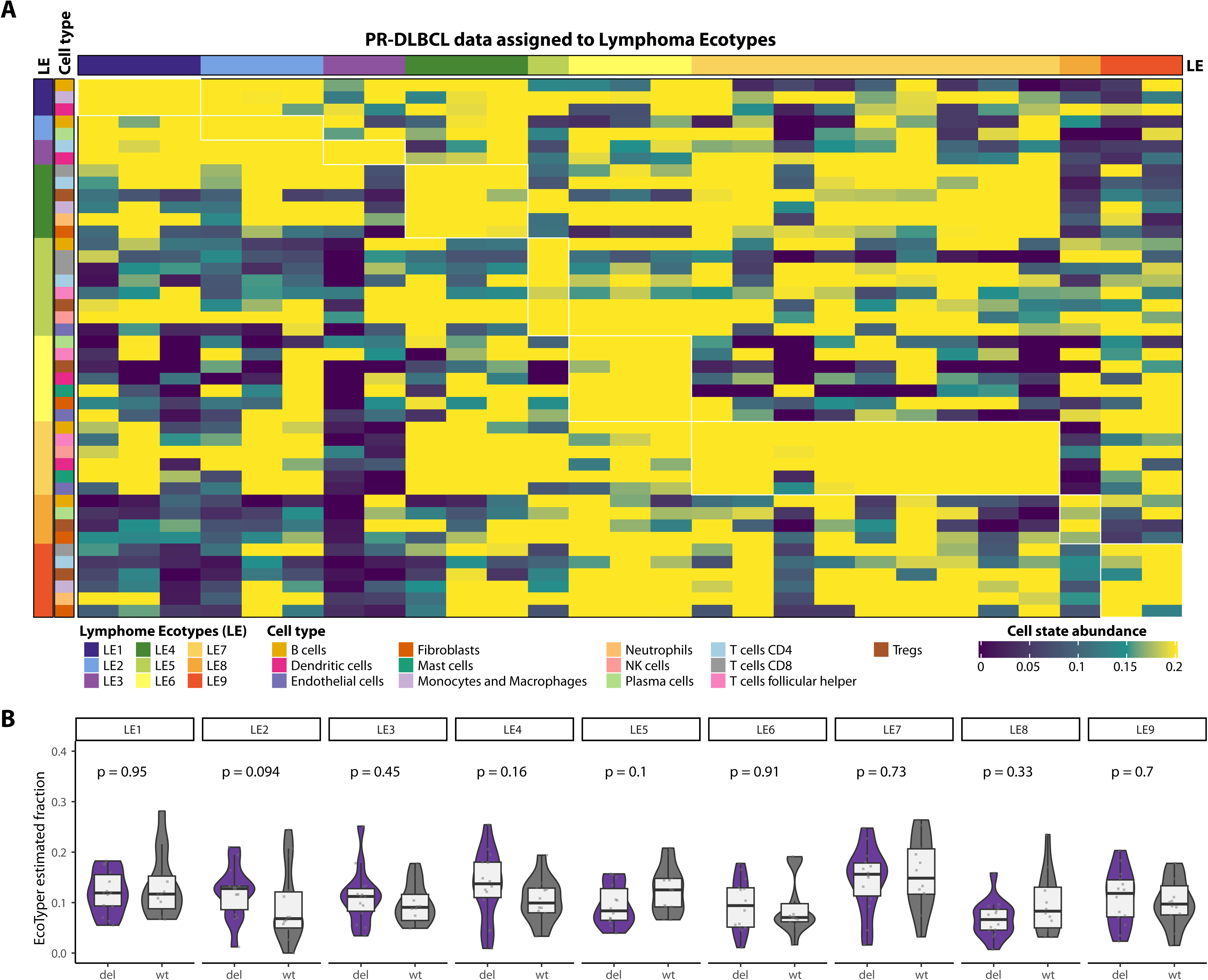
Deconvolution of PR-DLBCL transcriptomes employing the lymphoma EcoTyper. **A** Heatmap displaying the cell-type abundance as well as the predominant EcoType for a given PR-DLBCL sample. **B** Comparative analysis of lymphoma EcoType abundances according to the presence or absence of a large-scale deletion on chromosome 6 leading to strong MHC class I impairment as the most recurrent and striking feature in prDLBC, grouped by lymphoma EcoTypes.

## Discussion

Primary renal DLBCL poses a rare phenomenon associated with adverse clinical features and outcomes, including a so far unexplained CNS tropism leading to significant mortality, especially in relapsed and/or refractory cases ^6^. This study was initiated to unravel the molecular underpinnings of primary renal disease, to allow for an understanding as to why and how the disease originates in an organ without a genuine lymphatic niche, and to explain the clinically observed tendency of early CNS progression. To this end, we conducted an integrative analysis employing WES, RNA-seq and array-based SCNV analysis on the largest cohort of PR-DLBCL studied to date.

Hereby, we made the following noteworthy observations. First, we provide a detailed molecular landscape of PR-DLBCL and observe a strong degree of heterogeneity in terms of molecular clusters according to the LymphGen algorithm ^31^. Contrary to our initial expectation, we observed a relatively low frequency of *MYD88* mutations. Therefore only one case that was allocated to the characteristic MCD cluster, which is otherwise recurrently observed in both primary and secondary CNS lymphoma as well as testicular lymphoma. Tropism for immune-privileged sites appears to be driven by immune escape strategies, which may be augmented by activating mutations in the B-cell receptor signaling cascade ^31, 32, 44, 45^. Beyond this, we saw a high frequency of *SPEN* mutations previously described to be associated with early progression and inferior outcome in an all-comer DLBCL cohort ^46^. Significant deregulation of TSGs was further validated via an integrative analysis of SCNAs and transcriptional abundance. Among the genes most commonly affected by mutations within our cohort, potentially targetable vulnerabilities included recurrent mutations in genes like *EZH2* for which clinically active inhibitors have been described previously, including combinations with BCL2 inhibitors in patients with co-occurrence of *EZH2* and *BCL2* aberrations, which we observed in our cohort as well. At the same time, recurrent *CARD11* mutations, downstream of Bruton’s tyrosine kinase (BTK) may render therapeutic approaches, employing BTK inhibitors inefficient in a clinically meaningful subset of patients^47–49^. Given the peculiar tropism of PR-DLBCL, the abundance of *STAT3* mutations appears striking. In addition to the known cooperative interplay between *STAT3* and the NFkB signaling cascade in post-germinal DLBCL, this may further be explained as a selection advantage throughout clonal evolution. In keeping with previous observations that linked *STAT3* mutations and its elevated activity to the modulation of the abundance and function of regulatory T cells in the TME of solid cancers, this suggests a therapeutical perspective involving STAT3 inhibition which might be able to target the TME and its permissive impact on the immunological niche in PR-DLBCL ^50, 51^. An additional candidate driver for early progression and CNS dissemination is the relatively high rate of *MYC* aberrations in more than 17% of evaluable cases ^52^.

Second, we describe a profile of SCNA and mutations leading to significant impairment of MHC class I/II families and its immediately associated genes in approx. 95% of cases in the cohort, which significantly exceeds observations in all-comer DLBCL cohorts ^32, 53^. From these observations, we deduce a strong overlap between PR-DLBCL and large B-cell lymphomas of immune-privileged sites (IP-LBCL) regarding SCNAs. At the same time, PR-DLBCL apparently lacks the characteristic mutational profile resulting in tonic B-cell receptor signaling usually associated with this newly described entity ^1, 54^. It is tempting to speculate on a theory of a first and second hit in the central nervous dissemination of PR-DLBCL acquiring the typical driver mutations in a second step, following the initial presence of strong immune-escape mechanisms ^55^. To investigate this in future studies, paired analyses of renal and CNS tumors will be required. Deleterious aberrations in MHC class I components and related genes constitute well-documented phenomena in DLBCL. However, the frequency at which this is observed in the present study, extending to highly recurrent alterations of MHC class II genes poses a significant enrichment ^32^. Moreover, this aligns with previous observations in primary CNS lymphoma and testicular lymphoma regarding deletions of the *HLA* and *CDKN2A* loci and extends well beyond previous reports in ABC-DLBCL by both immunohistochemistry and gene expression profiling ^41^. Further in keeping with pathogenetically similar trajectories between PR-DLBCL and other IP-LBCL, we observed 10% of cases harboring *ETV6* rearrangements previously reported as recurrent, oncogenic structural variants and drivers of a post-germinal phenotype in PCNSL ^36^.

Third, we describe a characteristic transcriptional profile in cases harboring MHC class I SCNAs as a newly described hallmark of primary renal DLBCL. It is predominantly characterized by a strong enrichment for interferon alpha and gamma responses, MYC targets, and JAK/STAT signaling and downstream NFkB activation via TNF-alpha signaling. All of these mechanisms are known drivers, especially of ABC-DLBCL. TNF signaling was recently shown to aberrantly remodel the fibroblastic reticular cell (FRC) network and contribute to a tolerogenic TME hindering therapeutic efficacy of immune-based therapies ^51,56^. Further, several metabolic processes including glycolysis, fatty acid metabolism, and oxidative phosphorylation appear particularly engaged. This could be particularly interesting in future targeted therapeutical approaches exploiting this dependence, as recently described^57^. In addition, we observed a trend towards enrichment of LE2- and LE4-high states in our predominantly ABC-like group of MHC class I deleted tumors, in keeping with previous observations from all-comer DLBCL cohorts where these lymphoma ecotypes were associated with inferior clinical outcomes ^43^. The trend towards more LE5-high tumors in the non-MHC class I deleted cases with its underlying cell states being more prevalent in healthy lymphoid tissues integrates seamlessly with this notion.

The study presents several limitations, inherent to the retrospective design. Despite this being the largest cohort of PR-DLBCL to date the sample size remains limited and samples for the study were collected over more than two decades. This results in significant heterogeneity in terms of first- and later-line treatment approaches and limited follow-up information. These factors prohibit a meaningful integrative analysis of molecular findings and clinical endpoints. In addition, there is a potential for cases with occult non-renal primary manifestations, which may have been missed by diagnosing physicians. However, considering that secondary renal manifestations typically occur in advanced-stage disease, the likelihood of a significant fraction of patients harboring secondary renal manifestations is relatively low. In summary, we present the largest molecular study on PR-DLBCL to date, untangle the genomic and transcriptional heterogeneity of the disease, and uncover evidence of significant pathogenetic overlap between PR-DLBCL and IP-LBCL. Our observations may inform future risk-adapted therapeutic approaches.

## Data Availability

All data produced in the present work are contained in the manuscript

## Declarations

### Ethics approval and consent to participate

This retrospective study was approved by the ethics committee of the University of Lübeck (reference-no 18-356) and conducted in accordance with the declaration of Helsinki. Patients have provided written informed consent regarding routine diagnostic and academic assessment, including genomic studies of their biopsy specimens alongside the transfer of their clinical data.

### Availability of data and material

Raw fastq files have been deposited in the European genome-phenome archive (EGA) under the accession number EGA50000000386. OncoScan Array data has been deposited in Gene Expression Omnibus (GEO) under accession number GSE270422.

### Author contributions

Study concept: NG, ACF, HM

Data collection: NG, AK, VvK, PL, MK, HW, VB, SS, KK, HM, SS, KS, AA, ACF

Data analysis and creation of figures and tables: AK, NG, ACF, HB, NvB,

Initial Draft of manuscript: NG.

Critical revision and approval of final version: all authors.

## Acknowledgements

The authors would like to thank Tanja Oeltermann for her skilled technical assistance. AK and HB acknowledge computational support from the OMICS compute cluster at the University of Lübeck.

