## Supplementary Materials and Methods for "Molecular profiling of primary renal diffuse large B-cell lymphoma unravels a proclivity for immune-privileged organ-tropism"

**Transcriptome Data Quantification, Fusion Detection, and Analysis**

RNA-seq data (fastq files) was processed using the rnaseq (v3.13.2) workflow provided by nfcore. Fastp trimmed reads were mapped against GRCh38 using salmon (v1.10.1) ^1^. Fusions were detected using rnafusion (v3.0.1; nfcore) against GRCh38 as reference genome; fastp was used to remove low-quality bases from the sequencing data and Arriba and Fusioncatcher were used as fusion callers ^2, 3^. Fusion calls were filtered and manually inspected to reduce the number of false-positive fusion events.

Differentially expressed genes were detected using DESeq2 (v1.44.0) after removing genes with less than 10 counts in the dataset ^4^. To reduce variance from *log_2_* fold-changes, a heavy-tailed Cauchy prior distribution for effect sizes was used (shrinkage of *log_2_* fold-changes) ^5^. Pathway enrichment analysis against HALLMARK gene sets (msigdf R package v2023.2) on DESeq2 estimated shrinked *log_2_* fold-changes was performed using gage (v2.54.0); results with a corrected p-value below 0.01 were considered significant.

Classification into ABC/GCB subgroups (cell of origin) was performed using expression profiles of previously described candidate genes for GCB (*ITPKB*, *MME*, *BCL6*, *MYBL1*, *DENND3*, *NEK6*, *LMO2*, *LRMP*, *SERPINA9*) and ABC (*SH3BP5*, *IRF4*, *PIM1*, *ENTPD1*, *BLNK*, *CCND2*, *ETV6*, *FUT8*, *BMF*, *IL16*, *PTPN1*) ^6^. Briefly, expression profiles (counts) were quantile normalized, *log_2_* transformed, and *z*-normalized across genes. For each subtype, a score was computed for each sample *i,* and gene *j* follows

$${ABCscore}_{i}= \frac{\sum_{j=1\ldots n} E_{ij}}{n}$$

$${GCBscore}_{i}= \frac{\sum_{j=1\ldots n} E_{ij}}{m}$$

*E_ij_* was defined as the expression of gene *j* in sample *i*, *n* was defined as the number of ABC genes with expression > 0, and *m* was defined as the number of GCB genes with expression > 0. The subtype score was defined as

$${RNASubtypeScore}_{i}= {ABCscore}_{i}- {GCBscore}_{i}$$

A sample with an *RNASubtypeScore* above 0.25 was considered as ABC-subtype, below -0.25 as GCB-subtype, respectively. Systematic identification of cell states and cellular communities from bulk RNA-seq data was performed by applying EcoTyper for diffuse large B cell lymphoma ^7^.

**Statistical analysis**

If not reported otherwise, statistical analysis was performed using R (v4.4.0) and p-values were corrected using Benjamini-Hochberg correction. The following R packages were used: Tidyverse (v2.0.0)^8^ for data handling and plotting; maftools (v2.20.0)^9^ to summarize, analyze, and visualize variant data; EnhancedVolcano (v1.22.0) (https://github.com/kevinblighe/EnhancedVolcano) plot volcano plots; ComplexHeatmap (v2.20.0) and pheatmap (v1.0.12) to draw heatmaps; ggpubr (v0.6.0) for box and violin-plots. MHC genes for heatmaps were retrieved from the gene ontology domain cellular component (MHC_CLASS_I_PROTEIN_COMPLEX, MHC_CLASS_II_PROTEIN_COMPLEX).

**Pseudonymization**

The processing of personal data for this study was performed pseudonymously by using a case ID. Due to pseudonymization data backtracking specific to the individual for non-members of the study group is nearly impossible. Only the initiators of the study (NG, AK) have access to a file that is separately stored (password-protected) containing the details on pseudonymization.

**Supplementary Figures**

**Supplementary Figure S1.** Graphical representation of the bioinformatics workflow for WES data processing.

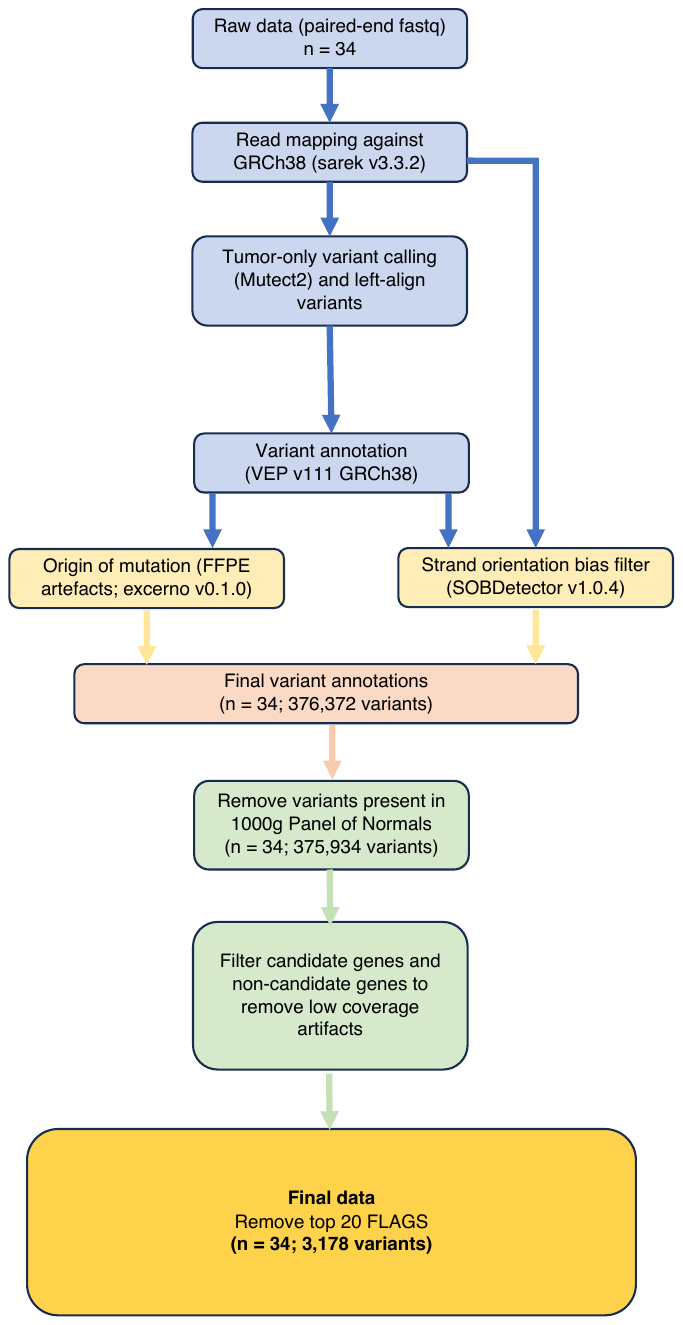

**Supplementary Figure S2.** Variants per variant class as well as variants per sample including information on variant subtypes.

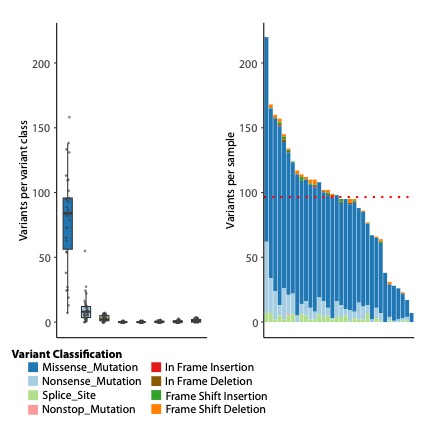

**Supplementary Figure S3.** Significantly mutated genes according to CoO according to Hans *et al.* (**A**) and BCL2 rearrangement status (**B**).

**Supplementary Figure S4.** Significant impact of SCNAs on gene-expression profiles shown exemplarily for TSGs

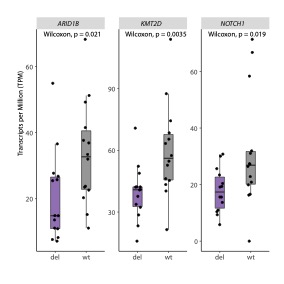

**Supplementary Tables:**

**Supplementary Table 1.** Baseline clinicopathological characteristics of the study cohort

**Supplementary Table 2.** Manually curated list of oncogenic fusions. fastp was used to remove low-quality bases from the sequencing data and Arriba and Fusioncatcher were used as fusion callers. Fusion calls were filtered and manually inspected to reduce the number of false-positive fusion events.

**Supplementary Table 3.** Mutations detected by WES in the present cohort of PR-DLBCL by type and frequency.

**Supplementary Table 4.** The genome-wide landscape of SCNA in 29 PR-DLBCL samples successfully analyzed on the Illumina OncoScan array following annotation via GISTIC (v2.0.22)

**Supplementary Table 5.** Annotation of individual cases by IHC, RNA-seq, and LymphGen.

**Supplementary Table 6.** Differential gene expression analysis according to the presence or absence of a large-scale deletion on chromosome 6 leading to significant impairment of the MHC class I apparatus in PR-DLBCL.

**Supplementary Table 7.** Gene-set enrichment according to the presence or absence of a large-scale deletion on chromosome 6 leading to significant impairment of the MHC class I apparatus in PR-DLBCL.

**Supplementary Table 8.** Distribution of lymphoma EcoTypes on individual cases of PR-DLBCL.
